# Occupational risk of COVID-19 by country of birth. A register-based study

**DOI:** 10.1101/2021.03.17.21253349

**Authors:** M Kjøllesdal, K Magnusson

## Abstract

**Aim:** To assess the role of occupation in the spread of COVID-19 among immigrants in Norway.

**Methods:** In 2.729.627 residents in Norway aged 20-70 years and born in Norway, Somalia, Pakistan, Iraq, Afghanistan and Turkey (mean [SD] age 44 (14) years and 51% men), we examined whether persons employed as taxi drivers, bus- and tram drivers, child care workers, nurses, personal care workers in health, food service counter attendants, waiters/bartenders, cleaners and shop sale persons had a higher risk of COVID-19, from April 1^st^ 2020 to December 2^nd^ 2020, compared to 1) Norwegian-born in the same occupational group and 2) all others with the same birth country and aged 20-70 years, using logistic regressions.

**Results:** Within each of the included occupational groups, immigrants had a greatly increased odds of COVID-19 when compared to Norwegian-born (OR ∼ 1.66-12.72). However, immigrants working in the selected occupations had the same odds of COVID-19 as person with *same* birth country not having the same occupation (OR∼1). Exceptions were Somalian-, Afghani- and Iraqi personal care workers in health services who had an increased odds of COVID-19 (OR 1.61 (95% CI 1.31, 1.98), OR 1.46 (1.06, 2.02) and OR 1.40 (1.03, 1.91), respectively) compared to others from the same country.

**Conclusion:** Immigrants holding various occupations implying close contact with others did not have higher odds of notified infection than others with the same country of birth, except for health care workers. Our study indicates that occupation is not an important driver of the high rates of COVID-19 among immigrants from Somalia, Pakistan, Iraq, Afghanistan and Turkey.

## Introduction

The COVID-19 pandemic hits different parts of a population to a different extent and infection rates have often been higher among immigrants than among native inhabitants (1-3). Immigrants from Somalia, Pakistan, and Iraq have presented with particularly high notification rates in Norway and Sweden (1,2). In the UK and US, both ethnic minorities (4-6) and people in disadvantaged socioeconomic positions (5, 7) are at high risk of COVID-19. Although ethnic minorities often are overrepresented in disadvantaged socioeconomic positions, various and composite measures of socioeconomic position could only modestly explain the high risk of COVID-19 among ethnic minorities (7-9). The importance of socioeconomic position in the high risk of COVID-19 among immigrants in Norway is not assessed.

Occupation may be important in the spread of COVID-19, with people in occupations with much contact with other people and without possibility to work from home (10), as well as people in low-income occupations and with short term contracts, being at especially high risk. We have recently shown that bartenders, waiters, food service counter attendants, taxi drivers and travel stewards were at increased risk of COVID-19 infection in the second wave of the pandemic (11), shedding new light on the earliest reports on occupational risk of COVID-19 that were performed in Singapore, Sweden and the UK (9, 11, 12). In the UK, the occupational risk among non-White workers was higher than among White, both in essential and non-essential occupations, with non-White essential workers being at the highest risk (9). We do not know whether some groups of immigrants have suffered more COVID-19 because of their overrepresentation in occupations representing high risk of infection, or whether their higher rates of COVID-19 are due to other factors. Knowledge about the role of occupations in the disproportional burden of COVID-19 among immigrants could inform us whether actions targeting occupations at risk could be a means to facilitate a decline in infections among immigrants, or whether increased risk seems to be caused primarily by other factors associated to being immigrant.

Thus, using data covering all employees registered in Norway, we aimed to study whether immigrants from five countries of birth with a large population in Norway and with a high incidence rate of COVID-19 (Somalia, Pakistan, Iraq, Afghanistan and Turkey) employed as car, van- and taxi drivers, buss- and tram drivers, child care workers and teachers’ aides, nurses, personal care workers in health, food service counter attendants, waiters/bartenders, cleaners and shop sale persons had an increased risk of COVID-19 compared to Norwegian-born in the same occupational groups and compared to others with same country of birth. These occupations were chosen as they were commonly held in the included immigrant groups and had high rates of COVID-19 infection.

## Methods

The Norwegian Institute of Public Health (NIPH) established a new emergency preparedness register covering the entire population registered as residents in Norway in April 2020, as part of its legally mandated responsibilities (13). Physicians and laboratories are obligated to report all confirmed cases of COVID-19 to *Norwegian Surveillance System for Communicable Diseases (MSIS*) using standard case-based notification practices (14-16). In the emergency preparedness register, data from MSIS, the Norwegian patient register, the Norwegian population register and the Employer- and Employee register were compiled and linked at the individual level using the unique personal identification number provided everyone in Norway at birth or upon immigration. Data from MSIS are updated daily, whereas the Employer- and Employee register was updated August 25^th^ 2020, and the population register November 2020.

We focused on immigrants from the five countries with highest rates of notified COVID-19 infection in Norway; Somalia, Pakistan, Iraq, Afghanistan and Turkey. Our study population included everyone born in Norway, as well as everyone born in one of these five countries, who had a personal identification number in the Norwegian population register, living in Norway at March 1^st^ 2020, and between 20 and 70 years of age. Data of notified cases of COVID-19 were included from April 1^st^ (leaving out the very first weeks of the pandemic, when mainly health personnel and persons at risk were tested, and most cases were likely to be related to travel to skiing destination in Europe) to December 2^nd^ 2020 (11).

### Variables

Immigrant status was defined as being born outside Norway, with country of birth as registered in the population register. Consequently, children born in Norway to immigrant parents are not included among immigrants. Missing information on country of birth was set to having birth country Norway.

Occupation was reported in the Employer- and Employee register with a 7-digit code according to the Standard Classification of Occupation in Norway (STYRK-98) (17). We converted the codes to align with the Standard Classification of Occupations (ISCO-08 using the 4-digit codes (Norwegian STYRK-08), to allow for international comparisons (17, 18). We included employment conditions with number of contracted weekly working hour >1. Included occupations are described in Table 1.

**Table 1.** Occupations with high numbers of employed from selected immigrant groups and with high notification rates of COVID-19.

| Occupations | Code* |
| --- | --- |
| Car, taxi and van drivers (taxi drivers) | 8322 |
| Bus and tram drivers | 8331 |
| Child care workers and teachers' aides (child care workers) | 531 |
| Nurses | 2221+2223 |
| Personal care workers in health services (health care workers) | 532 |
| Food service counter attendant | 5246 |
| Waiters and bartenders (waiters) | 513 |
| Shop sale persons | 522 |
| Cleaners | 911 |

\*According to the International Standard for Classification of Occupation (ISCO-08 / STYRK-08). Names in brackets used from here.

### Statistical analyses

First, we calculated the rates of COVID-19 as (cases/population)*100.000 in groups by country of birth and by occupation (the latter given as per 1000 employees). Second, we assessed the association between occupation and COVID-19 in logistic regression analyses using two different reference categories: 1) Norwegian-born in the same occupational group, aged 20-70 years and 2) all others with the same birth country in the age group 20-70 years. This allow for two different comparison groups for each occupational group from each country: For example, the infection rate of taxi drivers born in Somalia is compared with the infection rate of 1) every Norwegian-born resident in Norway aged 20-70 who is a taxi driver and 2) every Somali-born resident in Norway aged 20-70 who is not a taxi driver. This approach allows us to shed light on whether the occupation plays a role in the increased risk of COVID-19 within the immigrant groups. All analyses were adjusted for age (linear variable) and sex. We performed sensitivity analyses by restricting the time period of included cases from 15^th^ of June (less restrictive testing regimen in Norway) to 2^nd^ of December 2020. We also performed sensitivity analyses comparing each occupational group (stratified by birth country) to all others in the age group 20-70 years with an occupation (excluding those with no occupation).

### Ethics

The study was approved by the Regional Ethics Committee South-East (REK 19864)

## Results

Our sample included 2.729.627 persons in the age group 20-70 years, of which 2.639.042 were born in Norway, 23.630 in Somalia, 19.048 in Pakistan, 21.179 in Iraq, 14.746 in Afghanistan, and 11.982 in Turkey. Mean age was 44 (SD 14) years, and 51% of participants were men. Of these a total of 24.4% were not registered with any occupation, 21.5% among Norwegian-born, and 35.2%, 45.1%, 45.5%, 47.1% and 50.3% among those born in Afghanistan, Iraq, Turkey, Pakistan and Somalia, respectively. The proportion of the foreign-born groups in each of the included occupational groups can be seen in Supplementary table A.

The occupations with the highest rates of notified cases of COVID-19 per 1000 varied with country of birth (Table 2). Among Somalis, the highest rates were found among taxi drivers, nurses and health care workers, whereas bus drivers, child care workers had the highest rates among Pakistanis (Table 2). Health care workers had the highest rates among Iraqis, bus drivers and cleaners among Afghanis and shop sale persons among Turks (Table 2). Among the selected occupations, food service counter attendants and waiters had the highest rate among Norwegian-born (Table 2).

**Table 2.**
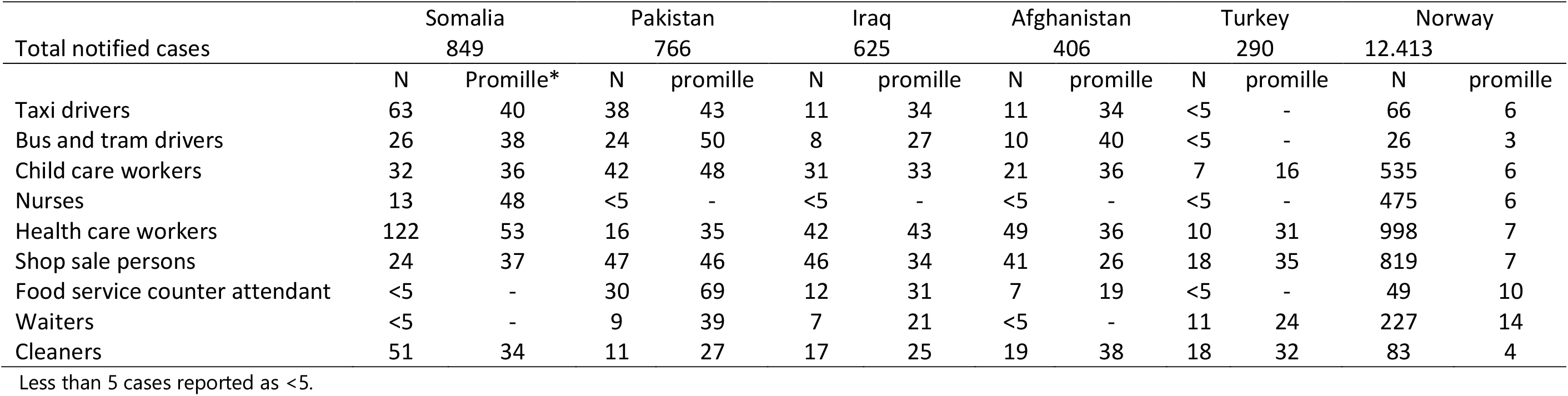
Number and rates per 1000 of notified COVID-19 in occupational groups by country of origin.

### Differences in occupational risk of COVID-19 between Norwegian-born and immigrants

Within the selected occupational groups, immigrants from Somalia, Pakistan, Iraq, Afghanistan and Turkey generally had higher odds of notified COVID-19 than Norwegian born (Table 4). The difference in odds between immigrants and Norwegian born within occupational groups often roughly reflected the difference in odds between immigrants and non-immigrants in general (Table 4). Among food service counter attendants and waiters, the differences in odds between immigrants and Norwegian-born were lower than the difference between groups in general (Table 3).

**Table 3.** Odds Ratio (95% confidence interval) of notified COVID-19. Reference: Norwegian-born in same occupational group. Age- and sex adjusted

|  | Somalia | Pakistan | Iraq | Afghanistan | Turkey | Immigrants total* |
| --- | --- | --- | --- | --- | --- | --- |
| Total | 6.89 (6.42, 7.40) | 9.33 (8.66, 10.05) | 6.11 (5.63, 6.63) | 4.74 (4.29, 5.25) | 5.45 (4.84, 6.13) | 6.65 (6.39, 6.93) |
| Taxi drivers | 5.94 (4.18, 8.46) | 7.76 (5.15, 11.69) | 5.63 (2.93, 10.80) | 4.15 (2.16, 7.98) | - | 6.56 (4.88, 8.86) |
| Bus and tram drivers | 8.38 (4.23, 16.60) | 11.31 (6.34, 20.21) | 5.92 (2.58, 13.56) | 8.98 (3.91, 20.61) | - | 12.72 (8.29, 19.55) |
| Child care workers | 4.88 (3.39, 7.03) | 9.60 (6.93, 13.29) | 5.53 (3.83, 8.00) | 5.18 (3.32, 8.08) | 2.85 (1.34, 6.05) | 5.81 (4.80, 7.05) |
| Nurses | 6.10 (3.46, 10.75) | - | - | - | - | 5.25 (3.37, 8.19) |
| Health care workers | 7.01 (5.76, 8.51) | 5.44 (3.29, 9.01) | 5.96 (4.34, 8.18) | 4.44 (3.30, 5.98) | 4.79 (2.54, 9.02) | 6.87 (5.95, 7.93) |
| Shop sale persons | 5.02 (3.31, 7.61) | 8.58 (6.33, 11.62) | 5.40 (3.99, 7.31) | 3.34 (2.42, 4.62) | 6.78 (4.20, 10.93) | 5.49 (4.66, 6.48) |
| Food service counter attendant | - | 10.47 (6.03, 18.16) | 3.85 (1.93, 7.66) | 1.73 (0.74, 4.04) | - | 4.12 (2.78, 6.09) |
| Waiters | - | 3.22 (1.61, 6.44) | 1.57 (0.73, 3.38) | - | 1.99 (1.06, 3.75) | 1.66 (1.14, 2.42) |
| Cleaners | 8.52 (5.86, 12.38) | 7.01 (3.62, 13.57) | 6.51 (3.75, 11.32) | 8.95 (5.17, 15.51) | 9.62 (5.64, 16.39) | 8.81 (6.66, 11.66) |

Regressions not performed in cells with <5 cases of notified COVID-19.
\*Somalia, Pakistan, Iraq, Afghanistan and Turkey together

**Table 4.** Odds Ratio (95% confidence interval) of notified COVID-19. Reference: all others aged 20-70 years from same country of birth. Age- and sex adjusted

|  | Somalia | Pakistan | Iraq | Afghanistan | Turkey | Immigrants total* | Norway |
| --- | --- | --- | --- | --- | --- | --- | --- |
| Taxi drivers | 1.20 (0.92,1.58) | 1.15 (0.82, 1.62) | 1.19 (0.65, 2.19) | 1.33 (0.72, 2.47) | - | **1.27 (1.06, 1.53)** | 1.23 (0.97, 1.57) |
| Bus and tram drivers | 1.10 (0.73, 1.65) | 1.31 (0.86, 2.01) | 0.96 (0.47, 1.96) | 1.47 (0.77, 2.82) | - | **1.28 (1.00, 1.62)** | 0.96 (0.66, 1.42) |
| Child care workers | 0.99 (0.69, 1.42) | 1.21 (0.87, 1.67) | 1.11 (0.76, 1.60) | 1.25 (0.80, 1.97) | 0.63 (0.30, 1.36) | 1.08 (0.90, 1.29) | **1.20 (1.10, 1.31)** |
| Nurses | 1.33 (0.75, 2.74) | - | - | - | - | 0.98 (0.63, 1.52) | **1.32 (1.20, 1.45)** |
| Health care workers | **1.61 (1.31, 1.98)** | 0.88 (0.53, 1.46) | **1.46 (1.06, 2.02)** | **1.40 (1.03, 1.91)** | 1.28 (0.67, 2.45) | **1.44 (1.25, 1.65)** | **1.33 (1.25, 1.42)** |
| Shop sale persons | 1.06 (0.70, 1.60) | 1.25 (0.92, 1.69) | 1.14 (0.83, 1.55) | 1.05 (0.75, 1.46) | 1.51 (0.92, 2.46) | 1.12 (0.95, 1.31) | **1.09 (1.01, 1.17)** |
| Food service counter attendant | 0.62 (0.15, 2.52) | **1.93 (1.32, 2.83)** | 1.04 (0.58, 1.87) | 0.80 (0.38, 1.72) | - | 1.28 (0.97, 1.68) | **1.45 (1.10, 1.93)** |
| Waiters a | - | 1.04 (0.53, 2.04) | 0.69 (0.42, 1.37) | - | 0.99 (0.54, 1.83) | 0.71 (0.50, 1.01) | **2.06 (1.81, 2.66)** |
| Cleaners | - | 0.65 (0.36, 1.20) | 0.85 (0.52, 1.38) | 1.37 (0.86, 2.19) | 1.36 (0.84, 2.21) | 0.97 (0.80, 1.17) | 0.82 (0.66, 1.10) |

Regressions not performed in cells with <5 cases of notified COVID-19
\*Somalia, Pakistan, Iraq, Afghanistan and Turkey together

In sensitivity analyses including only the period after mid-June 2020, the general difference between immigrants and non-immigrants remained.

### Occupational risk of COVID-19 within groups by birth country

Within groups of immigrants, there was little evidence of increased risk for COVID-19 associated with the occupations under study. However, health care workers had higher odds than others among persons born in Somalia, Afghanistan and Iraq, and waiters from Pakistan had higher odds of notified COVID-19 cases than other persons from Pakistan (Table 4). Among immigrants from these five countries in total, taxi drivers, bus- and tram drivers and health care workers had slightly higher odds than others for COVID-19. Among persons born in Norway, waiters, food service counter attendants, health care workers, nurses, childcare workers and shop sale persons had higher odds than others born in Norway for notified COVID-19.

In sensitivity analyses, excluding those with no registered occupation from the reference group, odds were slightly attenuated in most occupational groups (Supplementary table B).

## Discussion

In this explorative study of the relevance of occupation vs. birth country in contracting COVID-19, findings imply that occupation plays a very modest role in explaining high rates of COVID-19 among immigrants from Somalia, Pakistan, Iraq, Afghanistan and Turkey. For most occupations, immigrants from these countries had higher odds of COVID-19 than Norwegian born, and the difference in risk between immigrants and non-immigrants within occupational groups often reflected the difference between immigrants and non-immigrants in general. Among immigrants, only health care workers from Somalia, Afghanistan and Iraq and waiters from Pakistan had higher odds of COVID-19 than others with the same country of birth.

The study sheds new light on our recent study of COVID-19 infection rates in occupational groups (11), by showing differences in rates of notified cases by birth country within occupational groups. The findings resemble previous research, where socioeconomic factors could not explain ethnic differences in the burden of COVID-19 (7-9). This was also found in a Norwegian report where adjustment for occupation to a little degree attenuated differences in rates of COVID-19 among immigrant groups (19).

Many working people live in households together with others. Infections contracted at work might transfer to household members and close contacts. Immigrants may more often than others live in large or crowded households (18). This could imply attenuation of associations between occupation and COVID-19 in some groups of immigrants, as one occupation related case of COVID-19 possibly easier spread to several people in the same group. Assessing occupational risk in the context of (extended) families would be a next step analyses when data are available.

High odds of notified infection among health care workers and among nurses (seen only among Norwegian-born) could be related to frequent testing (11). In the beginning of the pandemic, only selected occupational groups, including nurses and other health personnel, as well as symptomatic persons returning from high incidence countries were offered a test. Later (after mid-June), anybody with symptoms of infection in the airways are urged to get tested. In sensitivity analyses excluding the period before 15^th^ of June, results did not change substantially, odds of COVID-19 were still higher among both non-immigrants and immigrants working as health care workers, whereas Norwegian-born nurses did not have higher odds compared to other Norwegian-born. Moreover, health personnel may have implemented better infection control measures as the pandemic progressed and may thus have lower rates of infection over time. Immigrants in general have not shown to have lower rates of testing for COVID-19 infection than Norwegian-born (20).

### Strengths and limitations

We use nation-wide register-based data on both notified COVID-19 and occupation. Because we include all residents of Norway aged 20 to 70 years and utilize routinely registered data, important strengths of our study are limited bias due to selection and misclassification. Important limitations are, first, we were not able to adjust for socioeconomic factors such as income or education, living conditions, urban living, health literacy or risky behavior. We acknowledge that all these factors are relevant for risk of infection and may be associated with persons’ occupation. Second, there may be differences within occupational groups in working contracts, working conditions and tasks. We hypothesize that workers on temporary contracts work more disadvantaged hours and perform tasks with higher risk of infection than others, as well as experience a stronger pressure to come to work even with symptoms of infection. Our data did not include information on whether participants had temporary or permanent position, but we suspect that immigrants more often than others work on temporary contracts. Third, other characteristics of occupations, such as distance to other people and possibility to work from home may (10) be just as relevant as occupations per se. A relatively large proportion of the population, about one in four, was not registered with any work, and excluding these from the reference group did not lead to substantial changes in results. A fourth limitation is that the numbers of immigrants with notified COVID-19 in each occupational group were quite small, especially among persons from Afghanistan and Turkey. Finally, we did not perform statistical analyses for groups with less than five cases, and there may also be uncertainties to estimates due to small samples and few cases also in other groups.

Persons born in Norway to immigrant parents are categorized as Norwegian-born, but as they have been raised in a home with immigrant parents, and many also in a society of people with the same country background, they may share many characteristics with immigrants. This could attenuate difference between Norwegian-born and immigrants, but as 3.5% of the Norwegian population are children of immigrants, and only 20% of them are above the age of 20 years (21), this should not impact our results notably. We assessed the association between occupation and COVID-19 infection in five immigrant groups in Norway, selected because they are relatively large groups with the highest rates of infection. Results may not be generalizable to all other groups of immigrants.

## Conclusion

Immigrants from Somalia, Pakistan, Iraq, Afghanistan and Turkey holding various occupations implying close contact with others did not have higher odds of notified infection than others with the same country of birth, except for health care workers. Thus, occupation did not seem to play an important role in the excess risk of COVID-19 infection among immigrants. Explanations for the high rate of infection is likely related to a complex set of factors including factors related to exposure and to underlying health.

## Data Availability

Data are from an emergency preparedness register established by the Norwegian Institute of Public Health (NIPH), as part of its legally mandated responsibilities. Data are now only available for NIPH employees.

## Acknowledgements

We would like to thank the Norwegian Directorate of Health, in particular Director for Health Registries Olav Isak Sjøflot and his department, for excellent cooperation in establishing the emergency preparedness register. We would also like to thank Gutorm Høgåsen, Ragnhild Tønnessen and Anja Elsrud Schou Lindman for their invaluable efforts in the work on the register. The interpretation and reporting of the data are the sole responsibility of the authors, and no endorsement by the register is intended or should be inferred. We want to acknowledge valuable input from Thor Indseth and Trude Arnesen to the manuscript.

## Author contribution

Marte Kjøllesdal performed the statistical analyses and the drafting of the article. Karin Magnusson contributed to the conception of analyses, in the interpretation of results and in the writing of the article. All authors gave final approval for the version to be submitted.

---

**Supplementary table A.** Number and proportions of workers within occupational groups by country of birth.

|  | Somalia<br>N=23.630 | Pakistan<br>N=19.048 | Iraq<br>N=21.179 | Afghanistan<br>N=14.746 | Turkey<br>N=11.982 | Norway<br>N=2.639.042 |
| --- | --- | --- | --- | --- | --- | --- |
|  | N (%*) | N (%) | N (%) | N (%) | N (%) | N (%) |
| Taxi drivers | 1562 (6.6) | 880 (4.6) | 327 (1.5) | 326 (2.2) | 220 (1.8) | 11169 (0.4) |
| Bus and tram drivers | 688 (2.9) | 478 (2.5) | 295 (1.4) | 250 (1.7) | 133 (1.1) | 8322 (0.3) |
| Child care workers | 891 (3.8) | 870 (4.6) | 928 (4.4) | 581 (3.9) | 435 (3.6) | 82477 (2.9) |
| Nurses | 270 (1.1) | 71 (0.4) | 113 (0.5) | 143 (1.0) | 54 (0.5) | 74600 (2.7) |
| Health care workers | 2297 (9.7) | 455 (2.4) | 980 (4.6) | 1354 (9.2) | 324 (2.7) | 145641 (5.2) |
| Shop sale persons | 652 (2.8) | 1025 (5.4) | 1349 (6.4) | 1584 (10.7) | 513 (4.3) | 125466 (4.5) |
| Food service counter<br>attendant | 91 (0.4) | 437 (2.3) | 388 (1.8) | 359 (2.4) | 115 (1.0) | 4942 (0.2) |
| Waiters | 105 (0.4) | 232 (1.2) | 334 (1.6) | 304 (2.1) | 463 (3.9) | 16567 (0.6) |
| Cleaners | 1519 (6.8) | 414 (2.2) | 677 (3.2) | 502 (3.4) | 559 (4.7) | 21688 (0.8) |

\*Proportion of groups by country of birth
Regressions not performed in cells with <5 cases of notified COVID-19.

**Supplementary table B.** Odds Ratio (95% confidence interval) of notified COVID-19. Reference: all others with any occupation from same country of birth (Those with no occupation excluded). Age- and sex adjusted

|  | Somalia | Pakistan | Iraq | Afghanistan | Turkey | Immigrants, total | Norway |
| --- | --- | --- | --- | --- | --- | --- | --- |
| Taxi drivers | 1.08 (0.81,1.44) | 1.09 (0.77, 1.56) | 1.04 (0.56, 1.92) | 1.23 (0.66, 2.30) | - | 1.16 (0.96, 1.40) | 1.17 (0.91, 1.49) |
| Bus and tram drivers | 0.99 (0.65, 1.49) | 1.26 (0.82, 1.94) | 0.85 (0.41, 1.74.) | 1.41 (0.73, 2.74) | - | 1.17 (0.91, 1.49) | 0.89 (0.60, 1.30) |
| Child care workers | 0.83 (0.57, 1.20) | 1.05 (0.74, 1.29) | 1.10 (0.75, 1.62) | 1.08 (0.68, 1.73) | 0.57 (0.26, 1.24) | 0.97 (0.80, 1.17) | **1.13 (1.03, 1.23)** |
| Nurses | 1.12 (0.63, 1.98) | - | - | - | - | 0.88 (0.57, 1.36) | **1.23 (1.12, 1.35)** |
| Health care workers | **1.44 (1.14, 1.83)** | 0.78 (0.46, 1.30) | **1.42 (1.02, 1.99)** | 1.22 (0.88, 1.68) | 1.20 (0.62, 2.31) | **1.31 (1.13, 1.51)** | **1.24 (1.16, 1.33)** |
| Shop sale persons | 0.93 (0.61, 1.42) | 1.12 (0.82, 1.53) | 1.05 (0.76, 1.44) | 0.90 (0.63, 1.25) | 1.42 (0.86, 2.35) | 0.99 (0.85, 1.17) | 1.03 (0.96, 1.11) |
| Food service counter attendant | - | **1.80 (1.23, 2.65)** | 0.94 (0.52, 1.68) | 0.73 (0.33, 1.51) | - | 1.15 (0.87, 1.17) | **1.39 (1.05, 1.85)** |
| Waiters | - | 0.96 (0.49, 1.88) | 0.61 (0.29, 1.30) | - | 0.91 (0.49, 1.69) | 0.64 (0.45, 0.91) | **1.98 (1.73, 2.26)** |
| Cleaners | 0.77 (0.57, 1.04) | 0.59 (0.32, 1.08) | 0.79 (0.48, 1.30) | 1.24 (0.77, 2.00) | 1.25 (0.76, 2.06) | 0.87 (0.72, 1.05) | 0.77 (0.62, 0.95) |

Regressions not performed in cells with <5 cases of notified COVID-19.

**Supplementary table C.** Odds Ratio (95% confidence interval) of notified COVID-19. Reference: all others from same country of birth. Age- and sex adjusted. Cases 16.06-02.12

|  | Somalia | Pakistan | Iraq | Afghanistan | Turkey | Immigrants, total | Norway |
| --- | --- | --- | --- | --- | --- | --- | --- |
| Taxi drivers | 1.11 (0.80,1.54) | 1.12 (0.77, 1.62) | 1.06 (0.54, 2.07) | 1.65 (0.89, 3.07) | - | 1.19 (0.96, 1.46) | 1.32 (1.03, 1.69) |
| Bus and tram drivers | 1.04 (0.64, 1.68) | 1.12 (0.69, 1.82) | 0.92 (0.43, 1.96) | 1.60 (0.80, 3.17) | - | 1.12 (0.85, 1.48) | 0.98 (0.65, 1.47) |
| Child care workers | 0.96 (0.62, 1.46) | 1.30 (0.92, 1.84) | 1.10 (0.74, 1.73) | 1.16 (0.69, 1.94) | 0.74 (0.35, 1.60) | 1.11 (0.91, 1.34) | 1.22 (1.11, 1.33) |
| Nurses | 1.23 (0.63, 2.41) | - | - | - | - | 0.91 (0.55, 1.50) | 0.98 (0.88, 1.10) |
| Health care workers | 1.35 (1.05, 1.73) | 0.84 (0.48, 1.48) | 1.41 (1.00, 1.99) | 1.29 (0.91, 1.83) | 1.04 (0.48, 2.23) | 1.24 (1.06, 1.46) | 1.25 (1.16, 1.34) |
| Shop sale persons | 1.29 (0.82, 2.01) | 1.18 (0.84, 1.66) | 1.14 (0.83, 1.58) | 1.08 (0.75, 1.56) | 1.71 (1.03, 2.83) | 1.19 (1.01, 1.41) | 1.15 (1.06, 1.23) |
| Food service counter attendant | - | 2.04 (1.37, 3.05) | 0.95 (0.50, 1.79) | 0.86 (0.38, 1.96) | - | 1.36 (1.02, 1.83) | 1.51 (1.13, 2.03) |
| Waiters | - | 1.23 (0.63, 2.42) | 0.65 (0.29, 1.46) | - | 0.97 (0.49, 1.91) | 0.79 (0.55, 1.15) | 2.11 (1.83, 2.42) |
| Cleaners | 1.07 (0.78, 1.46) | 0.55 (0.27, 1.12) | 0.88 (0.53, 1.45) | 1.21 (0.70, 2.09) | 1.34 (0.79, 2.27) | 0.97 (0.79, 1.19) | 0.83 (0.66, 1.04) |

Regressions not performed in cells with <5 cases of notified COVID-19.

**Supplementary table D.** Odds Ratio (95% confidence interval) of notified COVID-19. Reference: Norwegian-born in same occupational group (and total sample). Age-and sex adjusted. Cases 16.06-02.12

|  | Somalia | Pakistan | Iraq | Afghanistan | Turkey | Immigrants, total |
| --- | --- | --- | --- | --- | --- | --- |
| Total | 5.63 (5.19, 6.11) | 8.97 (8.28, 9.72) | 6.23 (5.,72, 6.78) | 4.28 (3.83, 4.79) | 5.15 (4.53, 5.86) | 6.13 (5.87, 6.41) |
| Taxi drivers | 4.25 (2.86, 6.32) | 6.98 (4.50, 10.31) | 4.90 (2.40, 9.99) | 4.42 (2.29, 8.51) | - | 5.36 (3.92, 7.33) |
| Bus and tram drivers | 5.60 (2.65, 11.81) | 9.56 (5.01, 18.24) | 5.71 (2.36, 13.78) | 8.27 (3.49, 19.61) | - | 10.70 (6.67, 17.16) |
| Child care workers | 3.83 (2.51, 5.86) | 9.62 (6.84, 13.54) | 5.54 (3.76, 8.16) | 4.33 (2.61, 7.18) | 3.18 (1.50, 6.76) | 5.39 (4.38, 6.64) |
| Nurses | 6.28 (3.19, 12.37) | - | - | - | - | 5.95 (3.58, 9.89) |
| Health care workers | 5.17 (4.09, 6.54) | 5.24 (3.01, 9.15) | 6.13 (4.34, 8.58) | 3.84 (2.74, 5.39) | 3.97 (1.87, 8.43) | 5.82 (4.93, 6.87) |
| Shop sale persons | 4.64 (2.98, 7.23) | 7.34 (5.25, 10.26) | 5.23 (3.81, 7.17) | 2.91 (2.05, 4.14) | 6.84 (4.18, 11.19) | 5.62 (4.21, 5.98) |
| Food service counter attendant | - | 10.13 (5.69, 18.01) | 3.34 (1.59, 7.01) | 1.58 (0.64, 3.91) | - | 3.80 (2.52, 5.73) |
| Waiters | - | 3.45 (1.72, 6.91) | 1.44 (0.63, 3.30) | - | 1.74 (0.86, 3.47) | 1.63 (1.10, 2.42) |
| Cleaners | 7.85 (5.25, 11.72) | 5.56 (2.60, 11.88) | 6.63 (3.74, 11.74) | 7.35 (3.95, 13.70) | 8.60 (4.82, 15.33) | 8.09 (5.99, 10.93) |

Regressions not performed in cells with <5 cases of notified COVID-19.

